# Operating Room Extubation After Cardiac Surgery: A Promising Practice or a Product of Selection Bias?

**DOI:** 10.1101/2025.09.22.25336347

**Authors:** Chelsea J. Messinger, Elizabeth Hall, Matthew R. Smith, Ariel Mueller, Min Hou, Jordan Bloom, Carolyn Mehaffey, Lauren Gibson

**Affiliations:** Department of Anesthesiology, Massachusetts General Hospital, Mass General Brigham, Boston, USA; Division of Cardiac Surgery, Massachusetts General Hospital, Mass General Brigham, Boston, USA

## Abstract

Extubation practices after cardiac surgery have increasingly shifted toward earlier strategies, including operating room extubation (ORE), although evidence regarding safety and outcomes remains mixed. On November 1, 2024, our center implemented a protocol requiring systematic consideration of ORE for all elective cases involving cardiopulmonary bypass, with prospective data collection through June 2025. Among 628 cases, 171 patients (27%) were extubated in the operating room, with a consistent monthly rate of approximately 30%. Patients had a median age 62 years and a relatively low burden of major comorbidities. Reintubation occurred in 5 patients (2.9%) and all patients survived to 30 days. Median intensive care unit stay was shorter for ORE patients compared with all elective cardiac surgery patients (25.5 vs 38.2 hours, respectively), while hospital stay was similar (5 days). These results likely reflect selection bias rather than causality. Nonetheless, they demonstrate the feasibility and safety of a standardized ORE protocol, supporting the need for a multicenter randomized trial to establish efficacy and define optimal patient populations.

## INTRODUCTION

Extubation practices following cardiac surgery have undergone substantial evolution in recent years. Traditionally, patients remained intubated postoperatively due to concerns regarding airway stability and respiratory complications. Advances in anesthetic techniques, physiologic monitoring, and perioperative care have since facilitated a shift toward earlier extubation—often within 6 hours of ICU admission—and, in select cases, immediate extubation in the operating room (ORE). ^1^

A small randomized clinical trial (N=100) conducted in 2018 demonstrated feasibility of ORE in elective, noncomplex cardiac surgery using cardiopulmonary bypass (CPB). ^2^ Several more recent, large retrospective studies have examined the efficacy of ORE. Some report reduced morbidity and shorter intensive care unit (ICU) length of stay compared with early ICU extubation. ^3–9^ In contrast, two large database analyses have identified associations between ORE and adverse outcomes, including higher rates of reintubation, longer operative times, and increased in-hospital mortality, though high-volume centers appear less affected. ^10,11^ These findings are limited by residual confounding given the substantial role of provider judgment in determining extubation candidacy—decisions often based on surgical and anesthetic factors not captured in electronic health records or national databases. This intractable confounding, combined with heterogeneity of observational studies, highlights the need for a pragmatic RCT.

As a preparatory step toward such a trial, our high-volume cardiac surgical center adopted a protocol in which all elective cardiac surgical patients are systematically considered for ORE. Here, we report preliminary safety and outcomes data from the first 8 months of implementation, with the aim of establishing feasibility ahead of a planned RCT of the protocol.

## MATERIALS AND METHODS

On November 1, 2024, we implemented a protocol requiring that potential for ORE be considered and discussed during the pre-procedural huddle (involving anesthesia, surgery, nursing, and perfusion teams) for all elective cardiac surgical cases undergoing CPB. Prior to this initiative, ORE was infrequent—occurring in approximately 3% of cases in 2024—and many anesthesiologists had limited experience with the practice.

Although all elective cases were considered, patients were identified as candidates for ORE during the huddle based on procedure complexity, age, body mass index, baseline mental status, airway considerations, and underlying pulmonary disease. The final decision to extubate at case conclusion rested on both pre-specified criteria and provider discretion. Relevant factors included airway stability, normothermia (core temperature ≥35 °C), hemostasis, CPB duration, metabolic status (ideally pH >7.30 and PaCO_2_ ≤50 mmHg), and other elements such as level of consciousness and adequacy of analgesia.

Data were prospectively collected from electronic medical records for patients extubated in the operating room between November 1, 2024, and June 30, 2025. Variables included demographic, preoperative, and intraoperative characteristics. Postoperative outcomes were obtained from the EMR and supplemented with standardized outcomes from the Society of Thoracic Surgeons (STS) Adult Cardiac Surgery Database. Monthly case volumes and length of stay were provided by the Department of Cardiac Surgery at MGH.

Descriptive statistics and baseline plots were generated in R (version 2025.05.1+513). Categorical variables are summarized as frequency (percentage), and continuous variables as median [Q1, Q3] based on visual inspection and quantitive

The use of these data for research was approved by the Mass General Brigham Institutional Review Board, which waived the need for informed consent.

## RESULTS

Of 628 elective cardiac surgical cases requiring CPB, 171 patients (27%) were extubated in the operating room. The monthly extubation rate averaged 29 (8.7)% (Figure 1A). Patients had a median age of 62 [54, 69] years and low prevalences of major comorbidities (Table 1). Median CPB and cross-clamp times were 127 [93, 172] and 92 [67, 124] minutes, respectively. Seven patients (4.1%) underwent deep hypothermic circulatory arrest (DHCA) and 7 (4.1%) underwent redo sternotomy. Aortic valve replacement, with or without concomitant aortic procedures, was the most common operation (30% of ORE patients) and had the highest extubation rate (73% of all AVR cases; Figure 2). Intraoperatively, patients received a median of 20 [20, 40] morphine milligram equivalents of hydromorphone and 72% of patients received midazolam before CPB. At CPB end, 60% of patients required ≤1 vasopressor/inotrope; at extubation this proportion increased to 82%. Fewer than 10% required ≥3 agents at either time point.

**Table 1.**
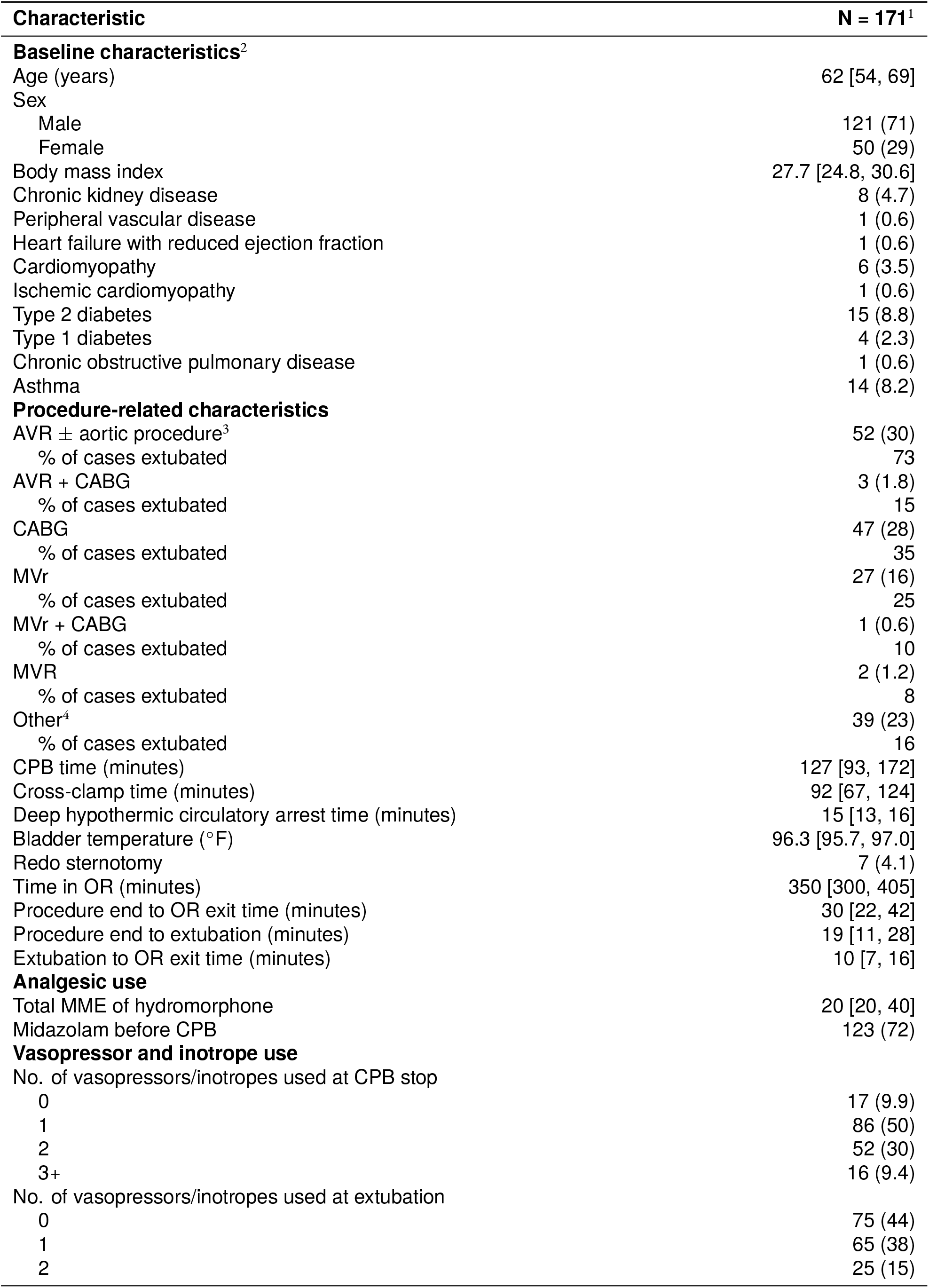
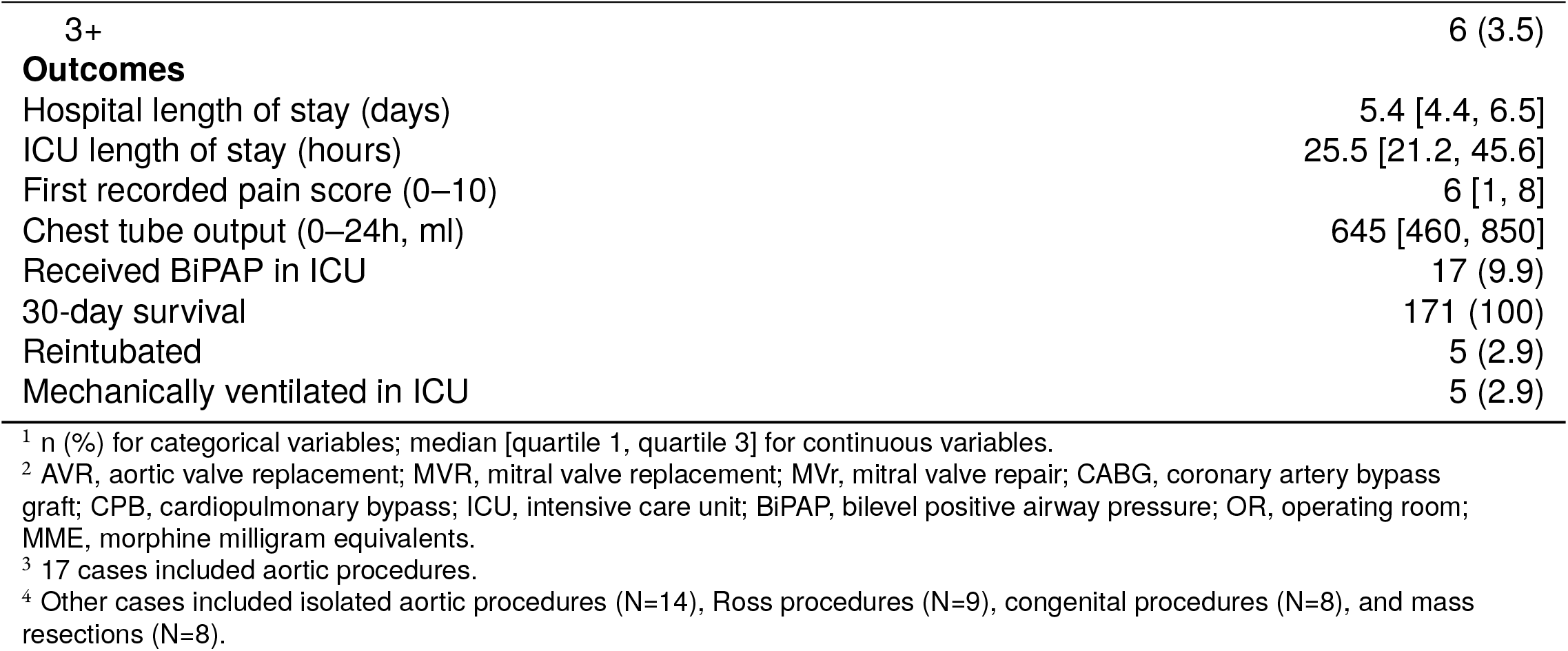
Pre-operative, intra-operative, and post-operative characteristics of patients extubated in the operating room (N=171).

**Table 2.** Reasons for and locations of reintubation among patients extubated in the operating room (OR). ICU, intensive care unit; DHCA, deep hypothermic circulatory arrest.

| Patient | Location | Reason |
| --- | --- | --- |
| A | OR | hypoxemia |
| B | OR | hypoxemia; pain requiring narcotics |
| C | OR | somnolent; obstructing airway |
| D | OR | poorly documented; given additional narcotic |
| E | ICU | hypothermia related; underwent DHCA |

**Figure 1:**
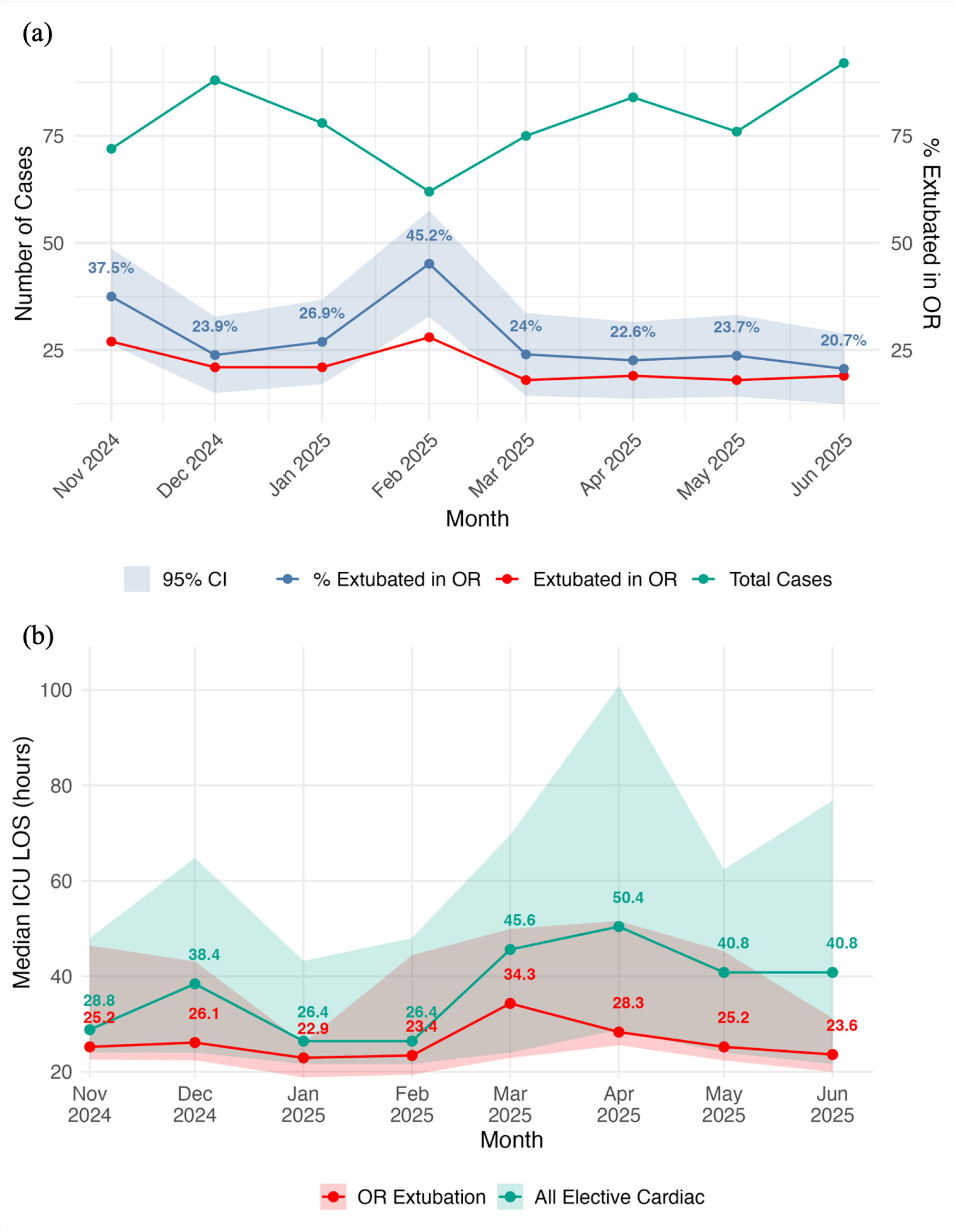
Operating room extubations and intensive care unit length of stay over time. (A) Number of total elective cardiac surgical cases, number extubated in the operating room (OR), and the proportion extubated in the OR over time. Ribbon represents 95% confidence interval (CI). (B) Median intensive care unit (ICU) length of stay (LOS) for patients extubated in the OR compared with all elective cardiac surgical patients. Ribbon represents interquartile range.

**Figure 2:**
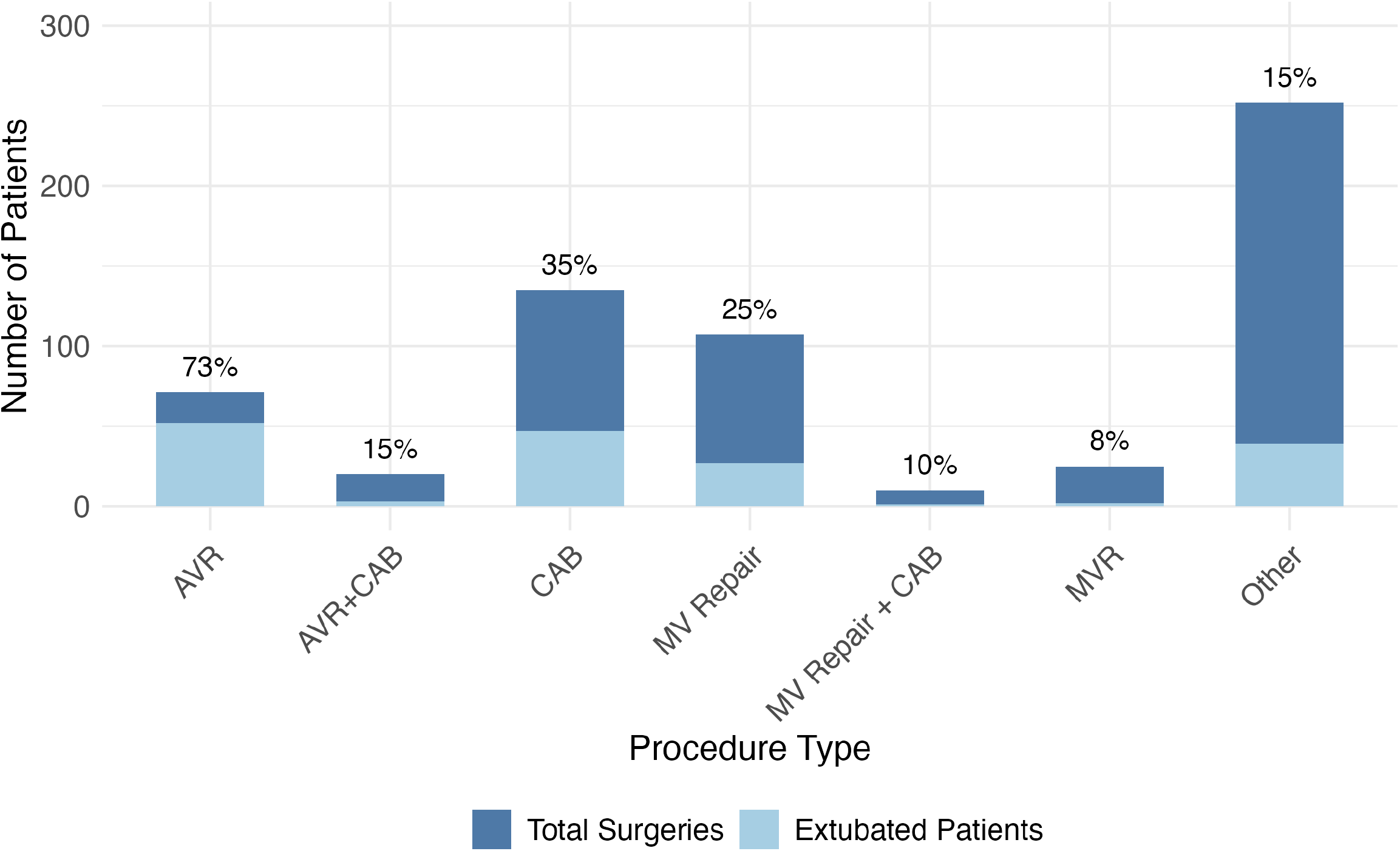
Number and percentage of patients for each procedure type who were extubated in the operating room. AVR, aortic valve replacement; CAB, coronary artery bypass; MV, mitral valve; MVR, mitral valve repair. Other cases included isolated aortic procedures (N=14), Ross procedures (N=9), congenital procedures (N=8), and mass resections (N=8).

The median interval from procedure end to OR exit was 30 [22–41] minutes. Five patients (2.9%) were reintubated; 4 in the OR and 1 in the ICU (Supplemental Table 1). Seventeen patients (9.9%) were placed on bilevel positive airway pressure (BiPAP) in the ICU, with use declining from 19% in November to 5% in June (Figure 3). All patients survived to 30 days.

**Figure 3:**
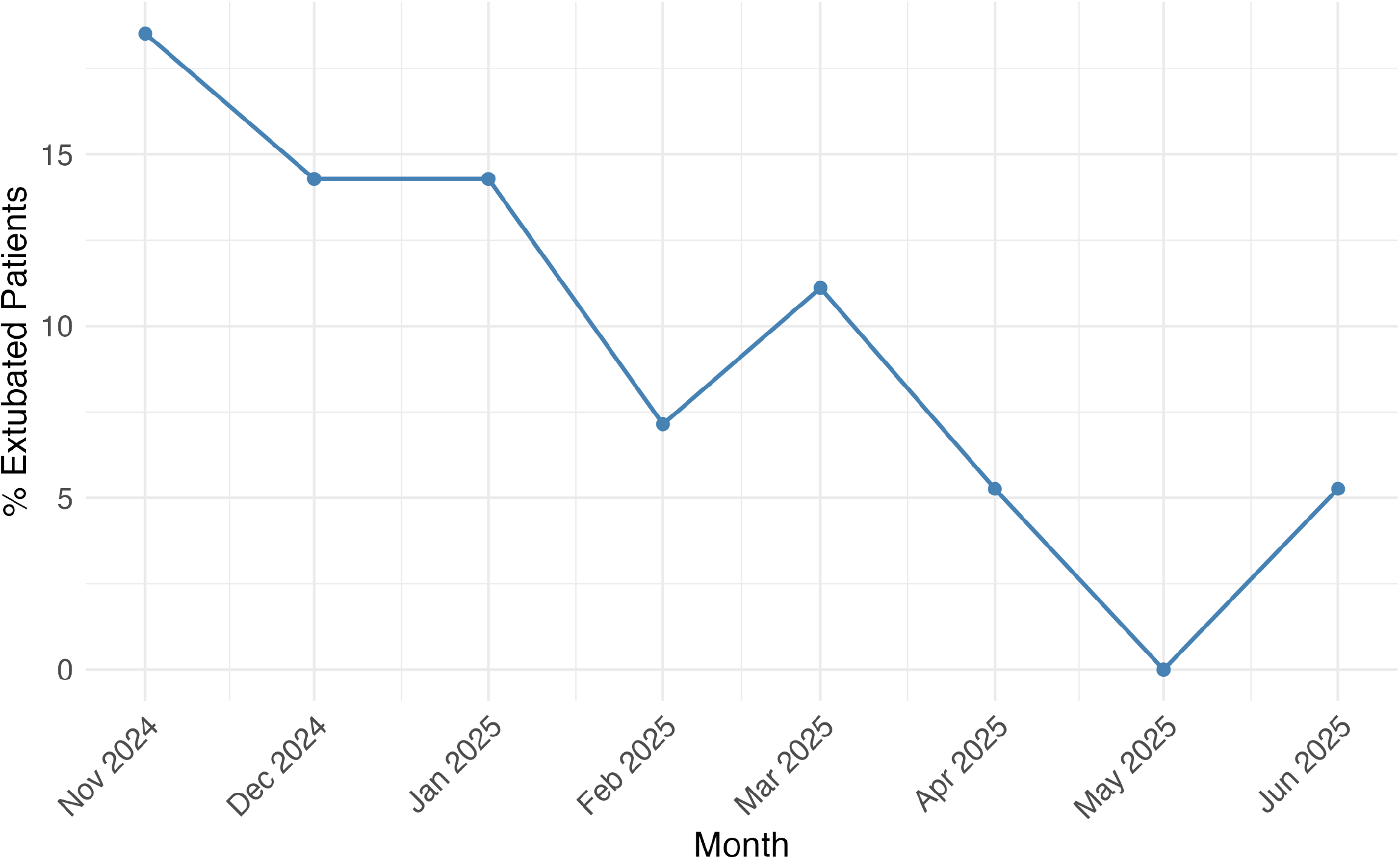
Proportion of patients extubated in the operating room who were placed on bilevel positive airway pressure (BiPAP) in the intensive care unit over time.

Median ICU length of stay was 25.5 [21.2, 45.6] hours for the ORE cohort versus 38.2 [23.3, 67.9] hours for the overall cohort, and median hospital length of stay was 5 [4, 7] days for both the ORE and overall cohorts (Figure 1B). ICU and hospital LOS among ORE patients varied slightly by procedure (Figure 4).

**Figure 4:**
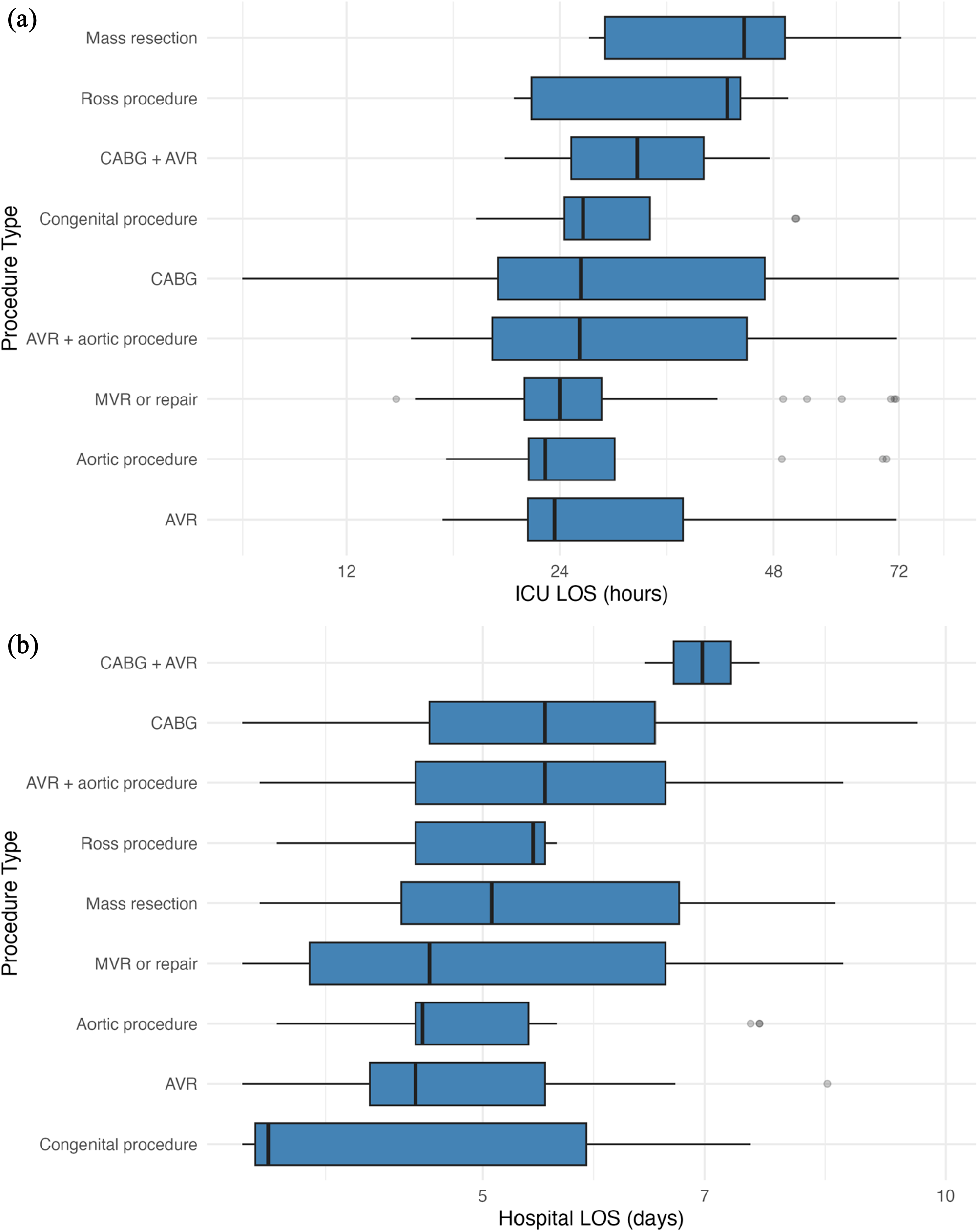
Box-and-whisker plots of length of stay (LOS) by procedure type. (A) Intensive care unit (ICU) LOS (hours) (B) Hospital LOS (days) Boxes show the interquartile range with medians marked; whiskers extend to 1.5 × the interquartile range, with more extreme values plotted as outliers. Axes use a log-like scale and are truncated at the 99th percentile to limit the influence of skewness and extreme values.

## DISCUSSION

Implementation of a protocol to consider ORE for all elective cardiac surgical patients was feasible, with a consistent monthly extubation rate of approximately 30% across a range of procedures, including complex operations such as Ross, DHCA, redo sternotomy, and congenital operations. Safety outcomes were favorable, with 0% 30-day mortality and a low reintubation rate (2.9%). BiPAP use in the ICU declined over time, suggesting improved patient selection and peri-extubation management.

Patients who were extubated in the OR were relatively young (median 62 years) and had low burden of major comorbid conditions (<10% for each), reflecting substantial provider discretion in patient selection. This subjectivity limits causal inference from retrospective comparative analyses, as many determinants of extubation candidacy are incompletely captured in clinical or national databases. The shorter ICU length of stay observed in this ORE cohort is therefore difficult to interpret given confounding by patient selection. Importantly, patient selection inherently reflects clinicaljudgement, and it remains essential that ORE be reserved for appropriate candidates, ideally within the framework of a well-defined protocol.

Future analyses will employ rigorous causal inference methods to assess the efficacy of our ORE protocol implementation, but these observational approaches remain inherently limited. A large, multi-center randomized clinical trial— already in active planning—is required to determine whether, and in which populations, ORE provides clinical benefit, with interim findings directly informing its design, patient selection, and implementation.

## Data Availability

All data produced in the present work are contained in the manuscript.

## AUTHOR CONTRIBUTIONS

Conceptualization: LG, CM, JB, EH, CJM; Methodology: CJM, MS, AM, MH; Investigation: CJM, EH, LG, CM, JB; Visualization: CJM; Writing: CJM; Editing: CJM, MRS, JB, LG; Supervision: LG, CM.

